# Long COVID: Assessment of Neuropsychiatric Symptoms in Children and Adolescents - A Clinical Data Analysis

**DOI:** 10.1101/2021.09.03.21257002

**Authors:** Jan Frölich, Tobias Banaschewski, Annabelle Ulmer

## Abstract

COVID-19 infections in adults often result in medical, neuropsychiatric, and unspecific symptoms, called Long COVID, and the premorbid functional status cannot be achieved. Regarding the course in children and adolescents, however, reliable data are not yet available.

**Objective:** 380 children and adolescents/young adults aged between 6 and 21 years, being treated for various psychiatric diseases in an outpatient clinical service, were examined for COVID-19 infections and Long COVID symptoms following a structured protocol.

**Results:** Three patients had COVID-19; one patient had symptoms of Long COVID in his medical history, but they could not be objectivized in an in-depth neuropsychiatric and neuropsychological assessment.

**Conclusions:** Long COVID seems to occur rarely in children and adolescents. Objectivizing the symptoms is a difficult task that requires various diagnostic considerations.

## Introduction

Since the beginning of the SARS-CoV-2 epidemic, there have been an increasing number of studies—especially involving adults—revealing that many of the affected patients enter a so-called Long COVID phase after the acute illness, a phase that can sometimes last for several months (Callard & Perego, 2021). These patients display a multitude of specific symptoms associated with medical diseases, but also neuropsychiatric and non-specific symptoms, and they fail to achieve the pre-infection functional level (Huang et al., 2021).

The term “Long COVID” is used when patients still show symptoms weeks to months after an infection with SARS-CoV-2. They have not recovered completely, they are the so-called “long haulers” (Baig, 2020). Thus far, the disorder has not been clearly defined in terms of nomenclature, specific disease criteria or a defined time period. Most patients rate themselves as moderately to severely impaired in their daily lives. Almost half of the patients meet the criteria for chronic fatigue syndrome (CFS).

Kedor et al. (2021) revealed in their observational study involving 42 adult patients (mean age 36.5 years) who had had COVID-19 that all patients without exception complained of fatigue six months after the acute infection, and nearly all had symptoms of post-exertional malaise (PEM), cognitive impairment and muscle pain.

Further studies indicate that seriously ill people were significantly more likely to experience long-term effects, including mental health issues. Old age, high BMI and history of lung disease were identified as predictive risk factors (Halpin et al., 2020). On the other hand, there are also reports of Long COVID experienced by individuals who were not seriously ill during acute infection, but nevertheless showed the apparently characteristic fatigue or tiredness after initial infection (Townsend et al., 2020). Females and individuals with a history of depression or anxiety disorder were more commonly affected.

Late neurological manifestations of a COVID-19 infection include meningoencephalitis, epilepsy or cerebrovascular stroke, and—much more frequently—the loss of the sense of smell and/or taste (German Society for Neurology, 2021; Walker et al., 2020).

There are very few reports of neuropsychiatric symptom constellations after the COVID-19 infection, even in adults. An Iranian study involving 201 patients with confirmed COVID-19 disease found that at least one neuropsychiatric symptom could be identified in 151 patients (i.e. 75.1 percent). Hierarchical clustering analysis yielded the following symptom groups: 1. anosmia and hypogeusia; 2. dizziness, headache, and reduced limb muscle strength 3. photophobia, changes in mental state, hallucination, vision and speech problems, seizure, stroke, and balance disturbance (Mirfazeli et al., 2020).

Children display much milder, less clinically pronounced symptoms of a COVID-19 infection, and severe courses of disease including death occur very rarely (Ludvigsson, 2020a; Bailey et al., 2020). It has repeatedly been reported that younger individuals have a significantly lower risk for developing coronavirus disease 2019 (COVID-19). Indeed a recently published study provides evidence that the airway immune cells of children are primed for virus sensing, resulting in a stronger early innate antiviral response to SARS-CoV-2 infection than in adults (Loske et al., 2021)

A case series with 5 children mentioned the following long-term effects identified thus far: fatigue, dyspnea, palpitations, chest pain, headache, concentration problems, muscle weakness, dizziness and sore throat, which are more severe in girls than in boys (Ludvigsson, 2020b).

Other than that, there are some publications on the long-term consequences of COVID-19 disease in children and adolescents. The first indications were reports from parents on social media. The internet platform “Long COVID Kids UK” (https://de.longcovidkids.org) was created by parents and launched in October 2020, offering the opportunity to those affected to exchange their experiences. Several 100,000 users were reached in an online message via Facebook and Twitter.

Based on these data Buonsenso et al. (2021) carried out the ‘Long COVID Kids Rapid Survey 2’, a follow-up study to record the frequency and type of symptoms. Inclusion criteria for enrollment into the study were a confirmed COVID-19 infection and symptoms for at least six weeks after the acute infection. Data were collected from 510 children (mean age 10.3 years, 56.3% girls). In 297 (58.2%) children, COVID-19 was confirmed by a PCR test (n = 141). COVID-19 was suspected, but not formally confirmed, in 209 (41%) children. 4.3% of the children received inpatient care, 74.1% were treated at home, and 12.2% remained asymptomatic. 43.7% of the children had no previous illnesses before they got infected. At the time of the survey, the respondents complained of the following symptoms: tiredness and weakness (87.1%), fatigue (80.4%), headache (78.6%), muscle pain (68.4%), post-exertional malaise (53.7%), irritability (51.4%) and dizziness (48%). 94.9% of the children surveyed complained of at least four symptoms. The most frequently mentioned changes to the status before the COVID-19 infection were changes in energy level (83.3%), changes in mood (58.8%), changes in sleep (56.3%) and changes in appetite (49.6%). 49.4% stated that they had repeatedly experienced significant improvements in their health with subsequent deterioration; only 10% stated that they had regained their pre-infection fitness level.

With regard to the deterioration in mental health and cognitive functions, the following results were obtained: poor concentration (60.6%), memory problems (45.9%), difficulty with everyday tasks (40%), and problems with working memory (32.2%).

In a prospective cohort study 518 children (≤ 18years old) admitted with confirmed Covid-19 to the hospital were assessed for persistent symptoms (>5 months). Median age was 10.4 years with 52.1% girls. At the time of the follow-up interview 126 (24.3%) participants reported persistent symptoms among which fatigue (53, 10.7%), sleep disturbance (36, 6.9%) and sensory problems (29, 5.6%) were the most common. Multiple symptoms were experienced by 44 (8.4%) participants (Osmanov et al., 2021).

## Clinical issue

The objective of this research was to assess the frequency and nature of possible neuropsychological/psychiatric symptoms after a COVID-19 infection using a sample of patients from the field of child and adolescent psychiatry.

## Methodology

### Standardized investigation algorithm (see Figure 1)

#### 1st stage of the investigation

444 patients and their parents, who were treated between January 1, 2021 and February 9, 2021 in a medical practice for child and adolescent psychiatry, were emailed an offer to take part in an investigation, defined in a standardized algorithm, for the presence of Long COVID symptoms as long as they were confirmed to have had a COVID-19 infection/disease. Patients in whom the infection was merely suspected, and there was neither laboratory proof nor a clinical diagnosis made by a physician, were explicitly excluded.

**Fig. 1:**
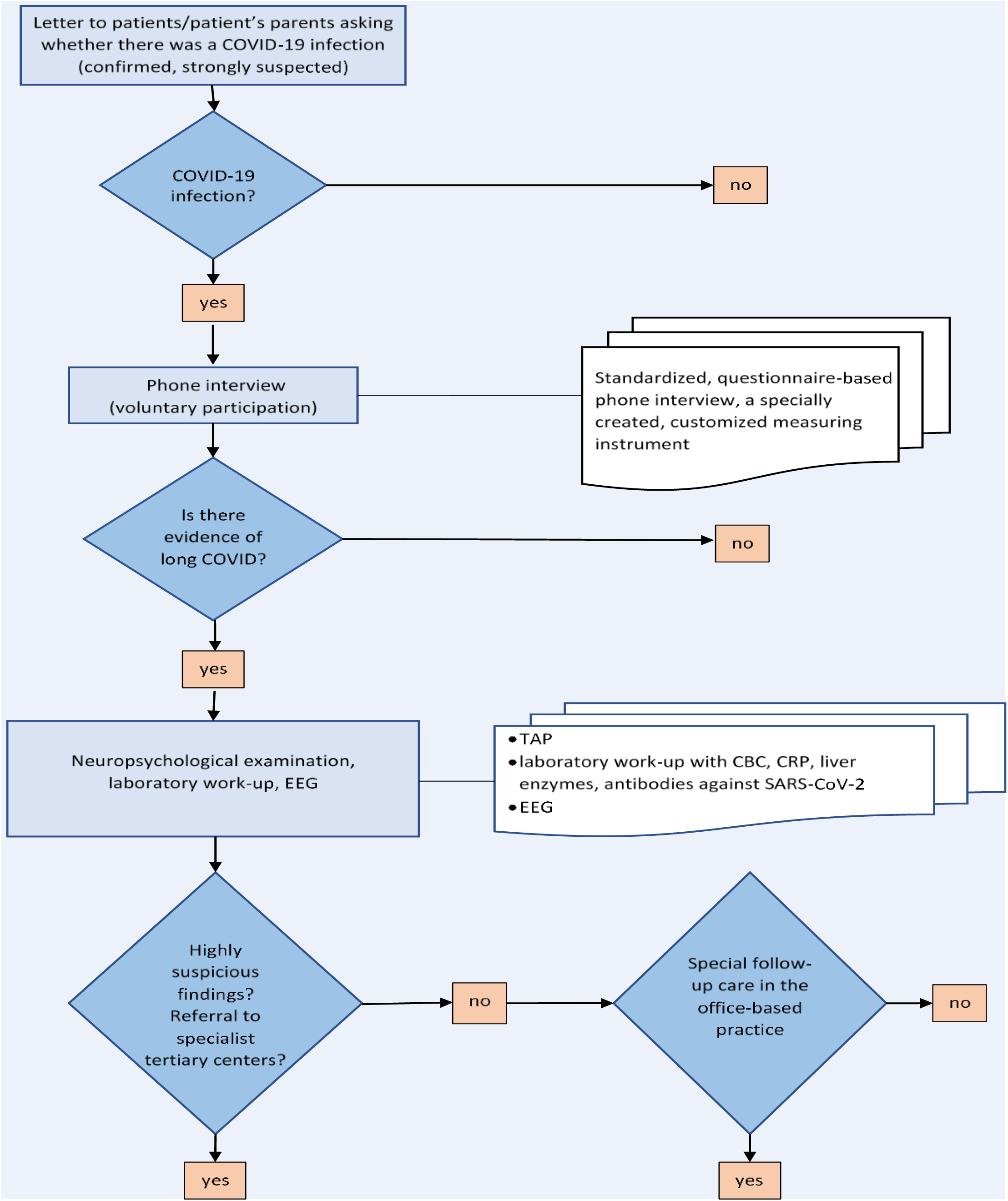
Standardized examination algorithm

The email contained a brief description of the symptoms of Long COVID and a detailed explanation of the investigation procedure. Feedback was requested from all patients either in writing, by telephone or during the following routine examination in our practice.

#### 2nd stage of investigation

Parents of the patients with a COVID-19 infection underwent a voluntary, standardized, questionnaire-based, 20-minute telephone interview conducted by a clinical psychologist. Since there are no standardized questionnaires for Long COVID for children and adolescents available, a measuring instrument was created specially for the purposes if this investigation. It is based on the neuropsychiatric symptoms most frequently reported in the literature (Rogers et al., 2020) (see Attachment).

#### 3rd stage of investigation

If the questionnaire revealed concrete red flags for the presence of Long COVID, a clinical-psychiatric and neuropsychological examination were carried out (test battery for Test for Attentional Performance (TAP) with the sub-tests Go/NoGo, divided attention, phasic alertness, cognitive flexibility) (Zimmermann & Finn, 2017). In addition, a panel of blood tests was performed including CBC, CRP, liver enzymes and antibody titer against SARS-CoV-2, as well as an awake EEG.

### Sample description

444 patients, aged 6–21 years (average age 14.0 years), 133 girls. Diagnoses: F90.0, F90.1, F90.8 n = 387; F84.5, F 84.8 n = 5; F32, F33 n = 29; F93 n = 6; F 42 n = 3; F95 n = 2; F60.31 n= 3; F41 n = 4; F63.9 n = 5.

All patients were receiving psychopharmacological treatment. Regular personal consultations take place during office hours, at least four times a year. For all patients, the initial evaluation followed a complete protocol including psychological testing, neuropsychological diagnostic work-up and EEG.

This was a non-interventional study. Informed parental consent was obtained from the three examined participants, and the study was performed according to the Ethical Principles for Medical Research Involving Human Subjects (Declaration of Helsinki, World Medical Association, WMA, 2013).”

## Results

Feedback was received from 380 patients either by email, by phone or in person during the following routine examination in our practice. Three patients (aged between 8 and 14 years; one girl and two boys) stated that they had COVID-19 diagnosed according to the above criteria.

During the interview, it was established that two patients had no complaints indicating Long COVID other than nonspecific symptoms due to psychosocial stressors caused by the pandemic.

In one patient with a F 90.0 diagnosis, however, there were specific indications. At the time of the examination, he was treated with a psychostimulant. No accompanying psychotherapy, only specialist care. Before the infection, he was well-controlled on his medication, enjoyed a stable psychosocial health, no psychiatric comorbidities, no history of somatic diseases.

The following clinical evidence emerged from the questionnaire-based interview: The patient stated that he had suffered cough, fever and a loss of the sense of taste during the acute infection. Overall, the course of the disease was rated as mild. Compared to his health status before the infection, at the time of the investigation for Long COVID ten weeks after the acute illness, he complained that the following symptoms had significantly worsened or had appeared: - *mild:* increased need for sleep, increased mood swings, speech/language abnormalities; - *moderate*: increased anxiety, concentration problems, depressed mood, states of confusion, increased aggressiveness, ideas of persecution and headaches; - *severe:* increased daytime fatigue

The interview revealed that there was no other life event during the same period that could explain these changes, and that the boy was generally able to cope with the psychosocial stressors of the pandemic. There were no newly diagnosed somatic diseases. The boy’s mother noted that the pre-infection symptoms of ADHD had worsened after the COVID-19 infection.

Psychiatric status: fully orientated to time, person and place; normal thought process and content, no memory impairment, no evidence of psychotic episodes.

Neurological status: Cranial nerves, reflexes, muscle strength and coordination unremarkable.

Neuropsychological examination under psychostimulant medication (TAP): normal values <u>for Go/NoGo</u> task, for divided attention and for alertness.

Awake EEG: Age-appropriate waveform without slowing, focal findings or hypersynchronous activity. There was only evidence of physiological, age-specific changes compared to the previous EEG.

Laboratory: negative COVID-19 serology; CRP, complete blood count with differential count and liver enzymes within normal limits.

## Discussion

According to the authors’ knowledge, this is the first standardized report of neuropsychological/psychiatric sequelae in children and adolescents carried out in the German-speaking countries according to defined and standardized selection and examination criteria.

All three patients stated that they continued to experience health issues and deficits after their COVID-19 infection. In the first step of the investigation, Long COVID was ruled out in two patients using a questionnaire-supported interview; rather, their health issues were attributed to the psychosocial restrictions of the pandemic. In the third patient, the interview revealed clear indications of Long COVID, but at the same time the reported complaints could not be confirmed in the clinical neuropsychological examination or with device-based investigations. In particular, antibodies were not detectable, although the diagnosis could be considered certain due to the specific symptoms (loss of the sense of taste) and given that the entire family was sick with COVID-19. At the same time, a negative anti-SARS-CoV-2 IgG antibody titer does not rule out past infection. According to a Chinese study, 40% of the examined patients with an asymptomatic infection, and 12.9% of the symptomatic patients, became negative for SARS-CoV 2 IgG antibodies again in the early convalescence phase (Long et al., 2020). A representative Israeli study (n = 2.391) identified a subgroup of symptomatic SARS-CoV-2 positive patients (∼ 5% of patients) who—regardless of age—remained seronegative, i.e. did not develop specific antibodies (Oved et al., 2020).

The results raise three fundamental questions with regard to Long COVID in children and adolescents:

1. Given the small number of studies conducted on this topic thus far, the incidence of Long COVID in this age group is one of the fundamental questions that need to be addressed.
2. Can specific, perhaps even age-typical, symptom constellations be identified, which first and foremost allow for a clear differentiation from the nonspecific symptoms that result from the indirect psychosocial stressors of the SARS-CoV-2 pandemic?
3. What valid examination instruments /methods help objectivize Long COVID based on the patients’ or their parents’ red-flag complaints acquired from medical history?

### Ad 1

In the examined sample of patients ranging in age from childhood to late adolescence, Long COVID seemed to be a rare event. The study by Buonsenso et al. (2021) investigating the frequency and severity of symptoms revealed that 90% of respondents had experienced symptoms. It should be emphasized, however, that this was an anonymous online survey and that there was no direct access to clinical data. Moreover, a COVID-19 infection could only be detected in two thirds of the respondents, and the data source was an online platform for patients with the symptoms of Long COVID.

With regard to this issue, an indirect comparison with the overall prevalence of SARS-CoV-2 infections in children and adolescents, which indicates that this age group is not only significantly less likely to be infected, but also less likely to become severely ill than adults, may make more sense. Debatin et al. (2020) examined 2,521 children aged 1–10 years and one parent of each child (n = 2,521) for current or past SARS-CoV-2 infection. The total study population comprised 5,042 subjects (2,521 children and 2,521 parents). The present preliminary analysis was carried out on 4,932 participants. Only one parent-child pair tested positive for SARS-CoV-2 RNA (0.04%) and they both had only mild symptoms. Overall, 19 children (0.8%; 95% CI, 0.5–1.2%) and 45 parents (1.8%; 95% CI, 1.3–2.4%) were seropositive. If it is postulated that COVID-19 is, in and of itself, a rare event in children and adolescents, it can be assumed that Long COVID—which by no means occurs in all infected patients—occurs even less often; in our study, it occurred in 0.26% of cases in three clearly established COVID-19 cases (0.79%). Naturally, this data should also be interpreted with some caution, since there was a high probability that there were undiagnosed cases in the study group due to a clinically covert or nonspecific course of infection. More recently other studies were published with regard to the prevalence of Long COVID in children: In a German study 1560 students (median age 15 years) grade 8-12 in fourteen secondary schools participated in the SchoolCovid19 study. Seroprevalence was assessed via serial SARS-CoV-2 antibody testing in all participants. Moreover, all participants were asked to complete a 12 question Long COVID survey regarding the occurrence and frequency of difficulties concentrating, memory loss, listlessness, headache, abdominal pain, myalgia/arthralgia, fatigue, insomnia, and mood (sadness, anger, happiness and tenseness). 1365 (88%) were seronegative, 188 (12%) were seropositive. Each symptom was present in at least 35% of the students within the last seven days before the survey but no statistical difference comparing the reported symptoms between seropositive students and seronegative students could be identified. The authors conclude that Long COVID might be less common than previously thought but possibly pandemic-associated symptoms regarding the well-being and mental health of young adolescents may be more prominent (Blankenburg et al., 2021).

In a large study in the UK with 258,790 children aged 5-17 years illness duration and symptom profiles were analysed for all children testing positive for SARS-CoV-2 for whom illness duration could be determined. Data from symptomatic children testing negative for SARS-CoV-2 were also assessed. 1,734 children had a positive SARS-CoV-2 test result. The commonest symptoms were headache (62.2%) and fatigue (55.0%). Median illness duration was six days (vs. three days in children testing negative). 4.4% children had illness duration ≥28 days (LC28), more commonly experienced by older vs. younger children. The commonest symptoms experienced by these children were fatigue (84%), headache (80%) and anosmia (80%). Only 25 (1.8%) of 1,379 children experienced symptoms for ≥56 days. Few children (15 children, 0.9%) in the negatively-tested cohort experienced prolonged symptom duration (Molteni et al., 2021).

### Ad 2

A standardized, stepwise diagnostic work-up of Long COVID performed in this study allowed for an adequate differentiation between the specific symptoms indicating Long COVID and the non-specific physical or psychological effects of the stressors of the pandemic. Moreover this differentiation is vital because the emotional stress caused by the illness, the persistence of complaints and the uncertainty about the further course of the disease could in and of itself be a cause for a depressed mood, anxiety or post-traumatic stress disorder (Del Rio et al., 2020). But also for reasons of cost containment, it is recommended that a targeted interview or questionnaire-based screening be performed prior to any comprehensive clinical diagnostic work-up. Currently, however, there is a complete lack of valid instruments for a solid preliminary diagnostic assessment.

### Ad 3

Despite a great deal of effort using clinical, neurological, neuropsychological, device-based examination modalities as well as serological tests, we were not able to objectivize the information gathered from medical history that included symptoms indicating Long COVID in the presented case because no health issues beyond the underlying disease came to light, neither in view of the current findings nor in comparison with the previous almost identical examination results.

It is therefore conceivable that a post-COVID syndrome will be diagnosed according to a set of patient complaint criteria to be formulated in the future, excluding any other somatic causes such as, for example, chronic fatigue syndrome (CFS) (Carruthers et al., 2011).

In the case of comorbid or previously existing (neuro-) psychiatric disorders such as ADHD, which our patient had, it can also be difficult to generate a meaningful differential diagnosis, since in ADHD there exist neuropsychological constellations of findings similar to those characterizing Long COVID (Nigg, 2005). It is also conceivable that stimulant medication for ADHD helps to significantly alleviate the daytime sleepiness typical of the post-COVID syndrome. Furthermore, neuropsychological examination findings may be influenced by the medication. Psychostimulants are used in accordance with the guidelines both in patients with narcolepsy (Mignot, 2012) and in patients with chronic fatigue syndrome (Blockmans &Persoons, 2016). Moreover, it should be kept in mind that the sleeping habits of children and adolescents changed during the pandemic and that a significant increase in daytime sleepiness was observed anyway (Bruni et al., 2021).

On the other hand, the results of our investigation can also be viewed as a call to develop more precise testing methods, which would be able to objectively verify symptoms gathered from medical history that are clearly suggestive of Long COVID.

The existing studies on the neuropsychiatric symptoms in adults actually used an even more extensive diagnostic work-up, incorporating a broad panel of tests. As an example, Mirfazeli et al. (2020) used a standard panel of blood tests including the complete blood count, CRP, blood gas analysis, liver enzymes, renal parameters, inflammatory markers, and creatine phosphokinase.

Kedor et al. (2021) used measuring instruments such as Chalder Fatigue Scale (Chalder et al., 1992) and Epworth Sleepiness Scale (Johns, 1991) to record fatigue and daytime sleepiness in their observational study. Since fatigue can be viewed as the cardinal symptom of Long COVID, its objective verification appears to be of utmost importance.

In the same study, a comprehensive laboratory analysis was also carried out that included CBC, lymphoycte subsets, IL-8 in erythrocytes, mannose binding lectin (MBL), CRP, immunoglobulins, ANA, ENA, C3/4, anti-TPO, TSH, fT3/4, ferritin, creatinine, liver enzymes, ACE, and NT-pro BNP. Furthermore, depending on the clinical symptoms, a lung and heart exam was carried out. Decreased MBL concentrations were found in 22% of the patients examined, and increased IL-8 concentrations in 43% of the patients.

Di Sante et al. (2021) recently documented significant immunologic differences between children that completely recovered from acute infection (n=17) and those with Long COVID (n=12). The group of Long COVID children showed significantly higher levels of plasmablasts, IgD-CD27+ memory and switched IgM-IgD-B cells. In contrast, healed children had significantly higher naÏve and unswitched IgM+IgD+ and IgM+CD27-CD38dim B cell subset. Moreover, IL6 and IL1β serum levels were elevated in Long COVID patients and consistently higher than children who had recovered after infection.

### The available neuropsychological research findings point in the following direction

Almeria et al. (2020) showed in their examined patients that neurological symptoms during the illness, diarrhea as well as hypoxia are associated with more substantial neuropsychological deficits with regard to working memory, attention, cognitive flexibility and phonetic fluency. The neuropsychological deficits can therefore be a direct consequence of brain damage as well as of prolonged reduction of oxygen saturation (Carda et al., 2020). Kumar et al. (2021) and Sozzi et al. (2020) highlight the importance of specific neuropsychological investigations in all patients complaining of cognitive, neurological and psychiatric problems over a longer period of time, on the one hand to initiate early interventions and on the other hand to differentiate a primarily virus-associated damage to the brain from that caused by secondary mental stress. Specifically, the authors mention an examination of executive functions, cognitive flexibility, learning and problem-solving abilities, attention and memory.

In summary, our research allows us to draw the following preliminary conclusions with greatest caution: Firstly, neuropsychiatric symptoms of Long COVID appear to be rare in children and adolescents. Secondly, there is a great need to obtain specific information on Long COVID symptoms for further diagnostic work-up with a reasonable expenditure of time using practical standardized questionnaires or interviews employed in the pediatric/child and adolescent psychiatry. Thirdly, the exent of the clinical diagnostic assessment to be carried out in such cases is rated as comprehensive and in-depth in terms of time investment and with regard to the necessary device-based investigations. It should should therefore be reserved for tertiary centers. An interdisciplinary approach appears to be indispensable.

## Data Availability

All data relevant to the manuscript are included in the article.

---

**QUESTIONNAIRE for Diagnostic Work-Up of Neuropsychiatric Symptoms After a Strongly Suspected or Confirmed COVID-19 Infection** (adapted from Rogers et al., 2020)

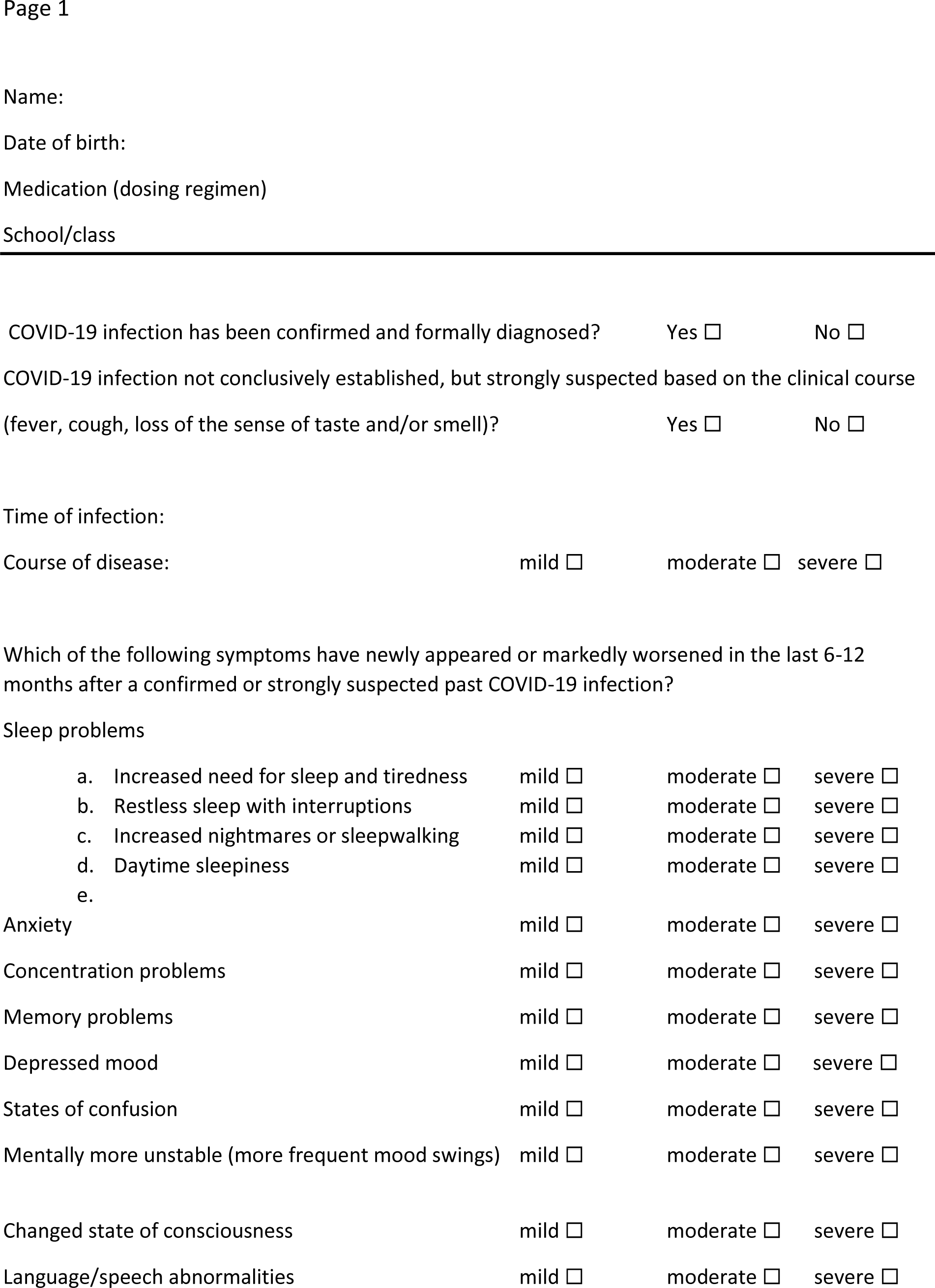
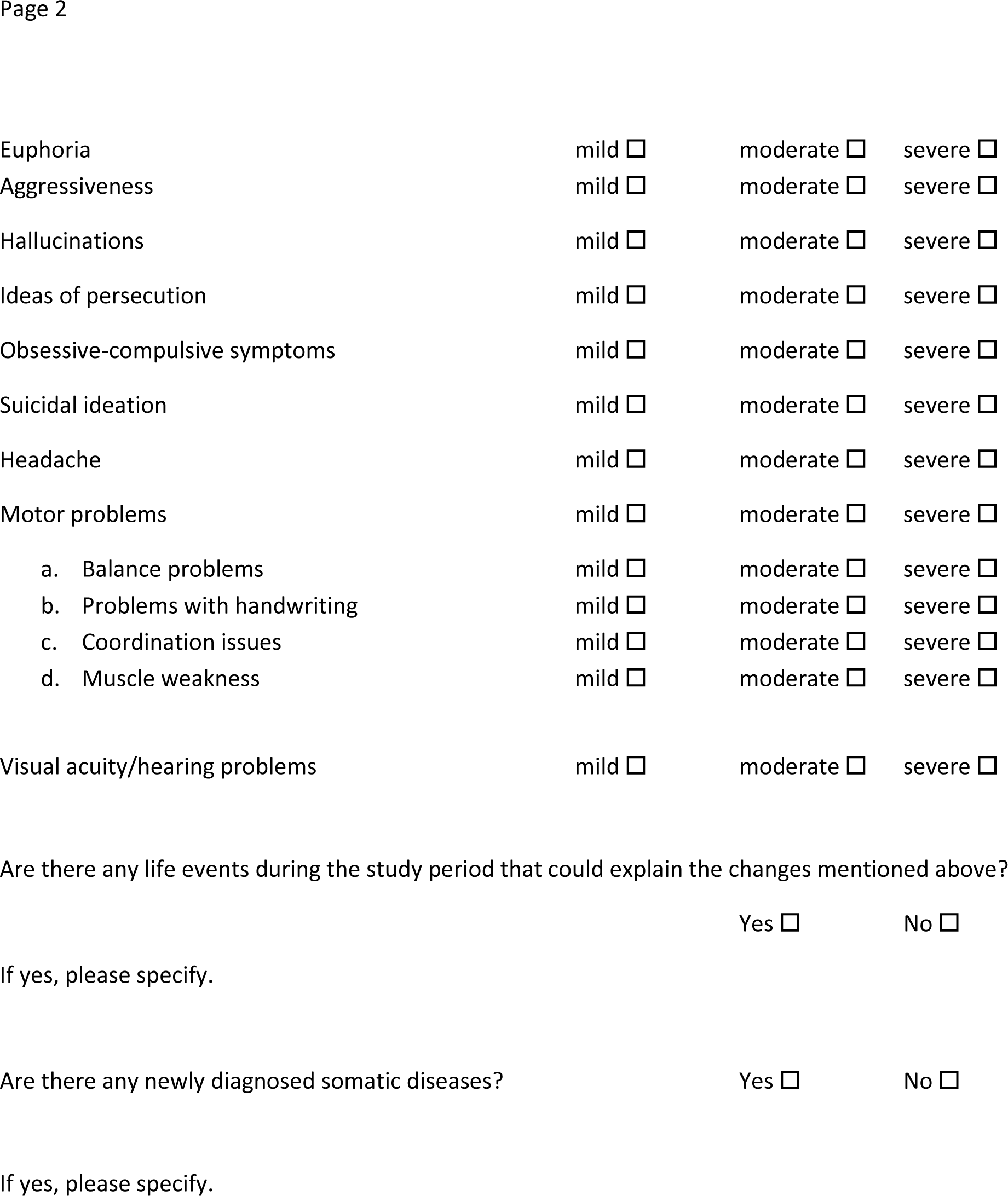

|  |  |  |  |
| --- | --- | --- | --- |
| a. Increased need for sleep and tiredness | mild ☐ | moderate ☐ | severe ☐ |
| b. Restless sleep with interruptions | mild ☐ | moderate ☐ | severe ☐ |
| c. Increased nightmares or sleepwalking | mild ☐ | moderate ☐ | severe ☐ |
| d. Daytime sleepiness | mild ☐ | moderate ☐ | severe ☐ |
| e. |  |  |  |
| Euphoria | mild ☐ | moderate ☐ | severe ☐ |
| Aggressiveness | mild ☐ | moderate ☐ | severe ☐ |
| Hallucinations | mild ☐ | moderate ☐ | severe ☐ |
| Ideas of persecution | mild ☐ | moderate ☐ | severe ☐ |
| Obsessive-compulsive symptoms | mild ☐ | moderate ☐ | severe ☐ |
| Suicidal ideation | mild ☐ | moderate ☐ | severe ☐ |
| Headache | mild ☐ | moderate ☐ | severe ☐ |
| Motor problems | mild ☐ | moderate ☐ | severe ☐ |
| a. Balance problems | mild ☐ | moderate ☐ | severe ☐ |
| b. Problems with handwriting | mild ☐ | moderate ☐ | severe ☐ |
| c. Coordination issues | mild ☐ | moderate ☐ | severe ☐ |
| d. Muscle weakness | mild ☐ | moderate ☐ | severe ☐ |
| Visual acuity/hearing problems | mild ☐ | moderate ☐ | severe ☐ |

Are there any life events during the study period that could explain the changes mentioned above?
Yes ☐ No ☐
If yes, please specify.
Are there any newly diagnosed somatic diseases?

